# Serum metabolomics identifies unique inflammatory signatures to distinguish rheumatoid arthritis responders and non-responders to TNF inhibitor therapy

**DOI:** 10.1101/2024.10.15.24315530

**Authors:** Michele Fresneda Alarcon, Yun Xu, Cassio Lima, Susanna Ford, Royston Goodacre, Marie M Phelan, Helen L Wright

**Affiliations:** Institute of Life Course and Medical Sciences, University of Liverpool, Liverpool, L7 8TX, UK; Centre for Metabolomics Research, Department of Biochemistry, Cell and Systems Biology, Institute of Systems Molecular and Integrative Biology, University of Liverpool, Liverpool, L69 7BE, UK; School of Life Sciences, University of Liverpool, Liverpool, L69 7BE, UK; High Field NMR Facility, Liverpool Shared Research Facilities University of Liverpool, Liverpool, L69 7TX, UK

**Author notes:** Correspondence: Dr Helen L Wright.

**Keywords:** metabolomics, serum, FTIR, rheumatoid arthritis, TNFi

## Abstract

**Introduction:** Rheumatoid arthritis (RA) is an auto-immune disease which causes irreversible damage to tissue and cartilage within synovial joints. Rapid diagnosis and treatment with disease-modifying therapies is essential to reduce inflammation and prevent joint destruction. RA is a heterogeneous disease, and many patients do not respond to front-line therapies, requiring escalation of treatment onto biologics, of which TNF inhibitors (TNF-i) are the most common.

**Objectives/Methods:** In this study we determined whether serum metabolomics, using nuclear magnetic resonance (NMR) and Fourier transform infrared (FTIR) spectroscopy, could discriminate RA blood sera from healthy human controls and whether serum metabolomics could be used to predict response or non-response to TNF inhibitor (TNF-i) therapy.

**Results:** NMR spectroscopy identified 35 metabolites in RA sera, with acetic acid being significantly lower in RA sera compared to healthy controls (HC, FDR<0.05). PLS-DA modelling identified 2-hydroxyisovalericacetic acid, acetoacetic acid, mobile lipids, alanine and leucine as important metabolites for discrimination of RA and HC sera by ^1^H NMR spectroscopy (averaged 83.1% balanced accuracy, VIP score >1). FTIR spectroscopy identified a significant difference between RA and HC sera in the 1000-1200 cm^-1^ spectral area, representing the mixed region of carbohydrates and nucleic acids (FDR<0.05). Sera from RA patients who responded to TNF-i were significantly different from TNF-i non-responder sera in the 1600-1700 cm^-1^ region (FDR<0.05).

**Conclusion:** We propose that NMR and FTIR serum metabolomics could be used as a diagnostic tool alongside current clinical parameters to diagnose RA and to predict whether someone with severe RA will respond to TNF-i.

## 1. Introduction

Rheumatoid Arthritis (RA) is a chronic autoimmune condition characterised by inflammation of synovial joints, causing swelling, stiffness and pain that can lead to irreversible joint damage if left untreated (Firestein 2003). Affecting 1% of the global population (Mizoguchi et al. 2018), it is the most common inflammatory joint disease (Cascao et al. 2010), and is a serious health and economic problem. In addition to joint inflammation, people with RA are at a higher risk of developing comorbidities such as osteoporosis, cardiovascular disease and lung disease (Figus et al. 2021), impacting negatively on quality of life and ability to work. Rapid diagnosis and initiation of preventative interventions for RA at the earliest stage can affect the disease course and halt progression of erosive disease. Recognising and diagnosing early RA from other inflammatory arthritides at the onset of disease is not straightforward and diagnosis is made from a spectrum of symptoms, inflammatory blood markers, x-ray and/or ultrasound of joints (Dey et al. 2023). We suggest that metabolomics techniques could help increase confidence in this diagnosis. Furthermore, predicting treatment outcomes has been a focus of many clinical studies (Anderson et al. 2020; Koo et al. 2021; Norvang et al. 2018). Despite its relative success, only around 60% of patients with severe RA have any measureable decrease in disease activity in response to TNF inhibitor (TNF-i) therapy, and only 30-40% achieve a low disease state, as classified by DAS28 scores (28-joint Disease Activity Score) below 2.6 (Kihara et al. 2017). Whilst more recent multiomics approaches have attempted to determine response to front-line methotrexate treatment (Wang et al. 2012) and TNF-i (Yoosuf et al. 2022) there is still currently no clinical diagnostic test to help stratify people with RA to an optimal treatment regime.

Serum metabolomics is a powerful tool for the differentiation of disease classes. Metabolic profiles are highly dynamic and differences may arise in disease even when protein or gene level do not show any change. NMR spectroscopy approaches have been used to obtain a global, unbiased views of small molecules in intact biofluids and tissues with minimal sample preparation, thus contributing to the understanding of the molecular characteristics of many diseases without the separation processes that may contribute significantly to the analytical variability (Gowda et al. 2014). Serum has been widely used in human blood metabolomics as it provides a wealth of metabolic information on health and disease, and is relatively easy to obtain by venipuncture without the need for invasive joint or tissue biopsy (Dunn et al. 2011). However, serum can often encounter potential confounding effects of haemolysis which affect the overall analysis (Gowda et al. 2014). In a recent study, serum was analysed by ^1^H-NMR to identify the markers in serum for early and reliable differential diagnosis of reactive arthritis and RA (Dubey et al. 2019). This work demonstrated that people with reactive arthritis are clearly distinguishable from controls and furthermore these patients can also be distinguished from people with RA based on the metabolic profiles.

Analysis of serum samples by ^1^H-NMR metabolomics requires minimal sample preparation, however the technology is not readily available in hospital laboratories. Developing an affordable, sensitive, specific and user-friendly portable diagnostic tool is critical to assure fast and accurate diagnosis of conditions. Fourier transform infrared (FTIR) spectroscopy is an analytical technique that has been widely used to understand composition of soft biological mediums and complex biofluids. Vibrational spectroscopy has seen an expansion in biomedical research for the evaluation of blood and blood derivatives such as plasma and serum (Crocco et al. 2023). Metabolic changes in body fluids alters the constituent molecules, providing strong guidance for subsequent clinical assessment (Baker et al. 2016). Attenuated total reflectance (ATR) FTIR serum analysis of patients diagnosed with breast cancer demonstrated high sensitivity and specificity in detection of the disease, close to those of mammography and ultrasound (Sitnikova et al. 2020). ATR-FTIR has also been shown to have a high sensitivity in the diagnosis of osteoarthritis in equine serum (Paraskevaidi et al. 2017). The presence of diagnostic autoantibodies such as anti-citrullinated protein antibody, rheumatoid factor, anti-neutrophil cytoplasmic antibodies, and anti-nuclear antibodies in auto-immune sera has been shown to correlate significantly with the wave numbers in the IR spectra (Durlik-Popinska et al. 2021).

The aim of this study was to use ^1^H NMR spectroscopy to investigate the metabolic composition of serum from people with RA. Using a combination of NMR and FTIR spectroscopy we then analysed the potential of serum metabolomics to discriminate RA sera from healthy individuals. Finally, we look to differentiate pre-treatment sera from responders and non-responders to TNF-i treatment based on follow up clinical data to determine potential spectral markers.

## 2. Methods

### 2.1 Ethical Approval

The study was approved by the NRES Committee North West (Greater Manchester West, Manchester, UK, Ref: 11/NW/0206) for the collection of sera from people with RA, and the University of Liverpool Central Research Ethics Committee (Ref: 1672) for the collection of sera from healthy controls (HC). All participants gave written, informed consent in accordance with the Declaration of Helsinki. All patients fulfilled the ACR criteria for the diagnosis of RA (Aletaha et al. 2010) and had severe RA with a DAS28 score >5.1. Response to TNFi was determined at 3 months by a decrease in DAS28 score >1.2.

### 2.2 Serum collection

Approximately 9 mL whole blood was drawn by venipuncture into Z serum clot activator monovette tubes (Griener Bio-one), followed by resting at room temperature (20-25°C) for 30 mins. The tube was then centrifuged at 1,500 x*g* for 15 min to retrieve the serum and 1 mL of sample was snap-frozen in liquid nitrogen and stored at -80°C.

### 2.3 ^1^H-NMR metabolomics

Immediately prior to NMR acquisition, the serum samples were defrosted on ice for 30 min. 120 μL of each sample was added to 120 μL of buffer master mix (200 mM sodium phosphate pH 7.4 in 20% ^2^H_2_O and 2.4 mM NaN_3_) and kept on ice throughout. The samples were centrifuged at 21,500 x*g* for 5 min at 4°C, and 200 μL of the supernatant transferred into a 3 mm outer diameter NMR tubes. Serum samples were analysed on a Bruker Ascend 700 MHz spectrometer (Massachusetts, USA) fitted with a 5mm TCI Cryoprobe, Avance III HD console and a SampleJet automated sample changer, keeping samples chilled (4-10°C) prior to acquisition. Prior to sample acquisition the spectrometer was calibrated for temperature and magnetic field stability using accepted quality assurance criteria as defined by Metabolomics Standards Initiative (MSI) (Sumner et al. 2007). The spectra for the serum samples were acquired at 37°C using ^1^H 1D CPMG edited pulse sequence (vendor supplied cpmgpr1d) was used to attenuate the signals from large molecules (proteins etc.) enabling quantitative appraisal of small molecule metabolites (Soininen et al. 2009). Each individual spectrum underwent quality control within the vendor supplied software (Bruker Topspin v3.6) by observing in each spectra water suppression quality, baseline correction and half height half width of the glucose anomeric peak at a chemical shift of 5.244 ppm. Alignment was checked by the anomeric glucose peak at 5.24 ppm and the binned (via integration of the area under each peak/bin region) using a pattern file previously produced for serum samples, with peaks annotated using the Chenomx NMR Suite 8.2 software (Chenomx Inc., Edmonton, Alberta, Canada). Representative metabolite peaks were selected using the correlation reliability score (CRS) developed by Grosman (Grosman 2020) which takes into consideration the property of NMR which states that peak intensities of correlated metabolites across the spectra will increase or decrease together.

### 2.4 Serum FTIR analysis

High throughput Fourier transform infrared (FTIR) spectroscopy was used to obtain infrared spectra from serum samples. Samples were first diluted in deionised water (1:5) in order to avoid signal saturation at the detector as well as non-linear effects. Samples were transferred to 96-well silicon substrates (Bruker Ltd, Coventry, UK) and dried (approximately 1 h) at 37°C prior to data collection. FTIR data were acquired in the mid-IR range (400-4000 cm^-1^), with 64 spectral co-adds and 4 cm^-1^ spectral resolution using a FTIR spectrometer (Bruker Invenio, Bruker Ltd, Coventry, UK). A total number of 4 infrared spectra were collected and from each sample/patient and all the spectra were vector normalised and baseline corrected. Mean average of the obtained spectra from each patient serum was calculated and carried forward for analysis.

### 2.5 Statistical analysis of metabolic profiles

The statistical calculations were carried out with software using R (v4.0.2) for univariate analysis and MATLAB 2023b (Mathworks, MA) for multivariate analysis. The comparison of NMR spectra between groups consisted of the analysis of the respective vectors of bucket integrals by using both univariate and multivariate statistics. Following tests for normality using the Shapiro-Wilks test, univariate analysis was carried out by Student’s t-test or Mann-Whitney test as appropriate, with application of a Benjamini-Hochberg false discovery rate (FDR) correction. For multivariate analysis, data were Pareto scaled before applying unsupervised PCA. Supervised PLS-DA was employed to build predictive models between experimental groups. The model performance was assessed by employing a five-fold double cross-validation (CV) procedure. This procedure employs two nested loops of CV: the outer loop and the inner loop. The data were firstly split into 5 folds (outer loop), each fold was used as a test set and the remaining 4 folds were used as the training set. The outer loop was repeated five times so that each fold had been used as a test set once. Within each training set defined by the outer loop, another five-fold CV (inner loop) was performed to tune the PLS-DA model (i.e., to select the optimal number of PLS-components with the minimal classification error). A PLS-DA model was then built on the training set with the optimal number of PLS-components and applied to the corresponding test set defined by the outer loop to estimated the performance of PLS-DA model which was assessed by balanced error rate (BER), defined as the average of the error rate of each class, and also the confusion matrix. In order to determine the statistical significance of the results and also to minimise the effect of variance caused by different combinations of training and test set, the double CV procedure was repeated 1,000 times with the order of samples randomly permuted each time, within each double CV procedure, a permutation test was also performed. Within each permutation test, the same double CV procedure was performed, but trained with the lables randomly permuted (i.e., a NULL model). The performance of the NULL was then compared with those of the model trained with correct labels (i.e., an observed model). Based on the results of 1,000 such comparisons, an empirical *p*-value was derived by calculating the percentage of the cases when the performance of the NULL model was better than that of the observed model, if there is not a single case that the NULL model was better than the corresponding observed model, the *p*-value would be designated as <0.001.

## 3. Results

### 3.1 ^1^H-NMR Serum Metabolomics analysis of RA sera

Serum spectra were divided into 160 bins and a total of 31 unique metabolites were annotated. CRS (Grosman 2020) was performed on the serum NMR data and a representative bin per metabolite was selected and carried forward for analysis (Table 1).

**Table 1:**
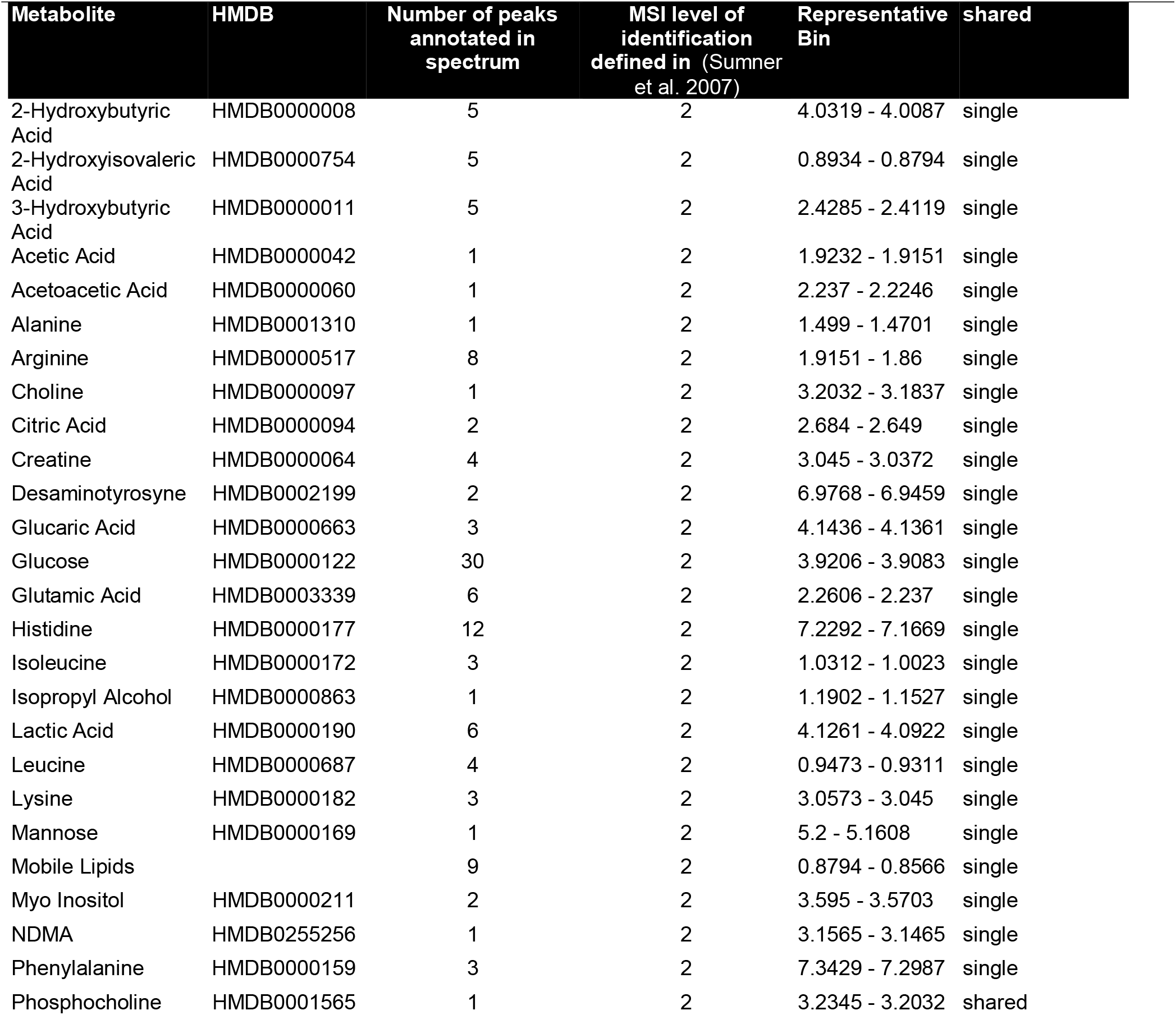
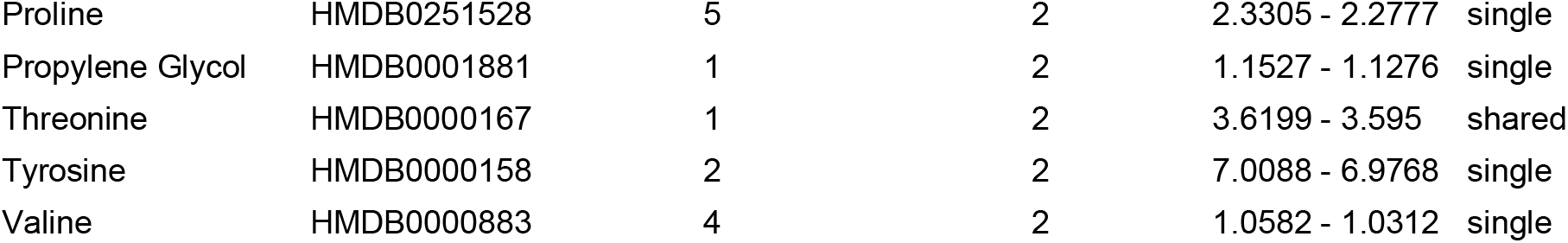
List of annotated metabolites from serum spectra. **Table** reports the total number of annotated peaks for each metabolite and the level of identification based on the metabolite reporting standards set out by the metabolomics society. A representative bin was chosen out of all related annotated peaks and brought forwards in the subsequent statistical analysis. HMDB = Human Metabolome Database.

Principal Component Analysis (PCA) did not highlight any separation between RA and HC groups (Figure 1A). Univariate analysis identified acetic acid has being lower in RA sera compared to HC (Figure 1B, FDR <0.05). The results of PLS-DA suggested that there is a partial separation between RA and HC samples. The averaged confusion matrix of the prediction of test sets is shown in Figure 1C, the averaged balanced accuracy was 83.1% with a *p* < 0.05. Although it is worth mentioning that the rather discrete NULL and observed distributions (Figure 1D) suggests that the statistical power was low and this is mainly due the number of biological replicate samples. The averaged VIP scores plot (Figure 1E) shows the importance of each variable to the model and the plot shows that 2-hydroxyisovaleric acetic acid, acetoacetic acid and mobile lipids were the most important metabolites for model prediction (VIP score >1), the latter representing high density lipids (HDL), low density lipids (LDL) and triglycerides (Figure 1E,F). Amino acids alanine and leucine were also important metabolites in the model prediction (Figure 1F).

**Figure 1:**
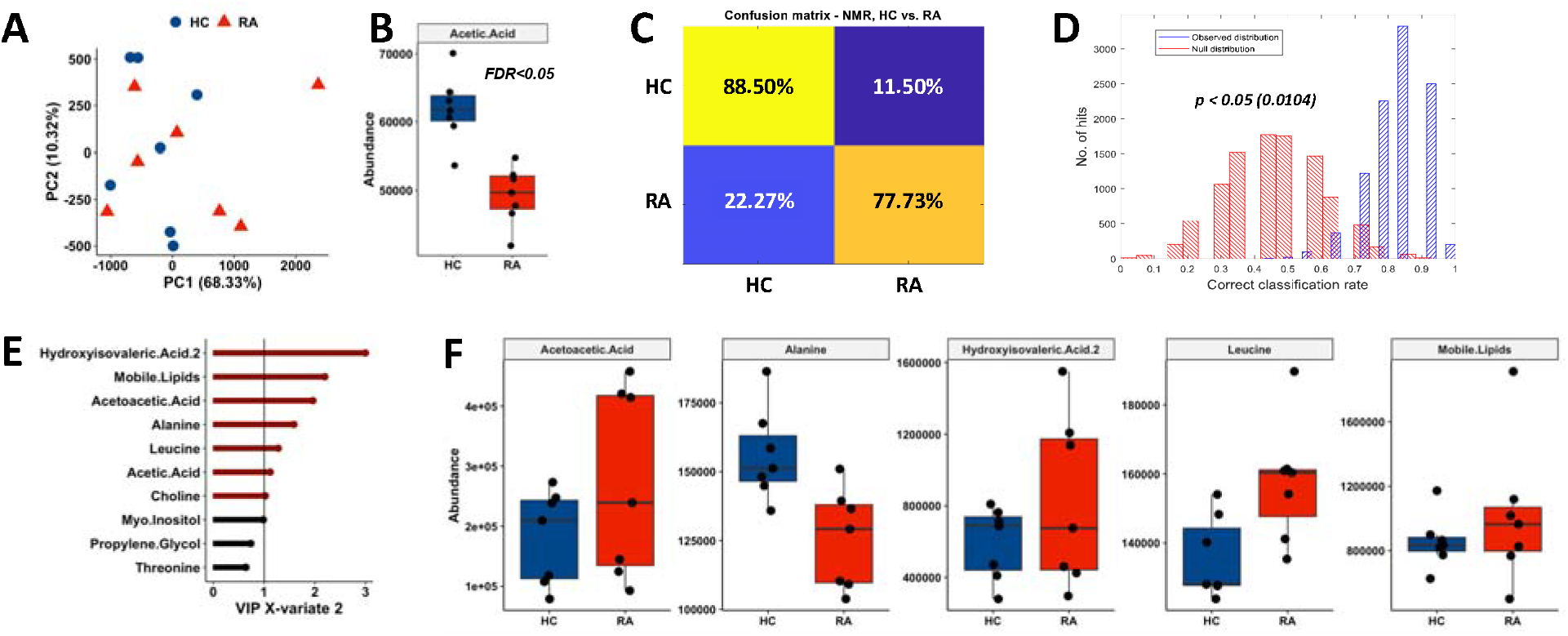
Univariate and multivariate analysis of RA and HC sera by 1H NMR metabolomics. (A) PCA scores plot of HC and RA serum. Brackets report the variance explained by the PC. (B) Acetic Acid was significantly lower in RA sera (FDR<0.05). (C) PLS-DA averaged confusion matrix of predicted tests sets. (D) The distributions of the performance of PLS-DA classification models: observed distribution (blue) and NULL distribution (red). (E) Top VIP scores of PLS-DA model built on disease discrimination. In red metabolites with VIP scores > 1. (F) Boxplots of important metabolites indicated by PLS-DA measured in HC and RA serum.

### 3.2 FTIR serum metabolomics

We next examined the chemical composition of serum samples derived from HC, RA and SLE using FTIR spectroscopy. Figure 2 shows the quantile plot of the whole serum spectra (400 – 4000 cm^-1^). The biomolecular and bonding vibration assignments are tentative and are based on numerous studies (Ghimire et al. 2020; Ramalingam et al. 2014) as shown in Supplementary Table 1. For the subsequent analysis the fingerprint region (940 – 1771 cm^-1^) was re-scaled by using extended multiplicate signal correction (EMSC) method (Afseth et al. 2012).

**Figure 2:**
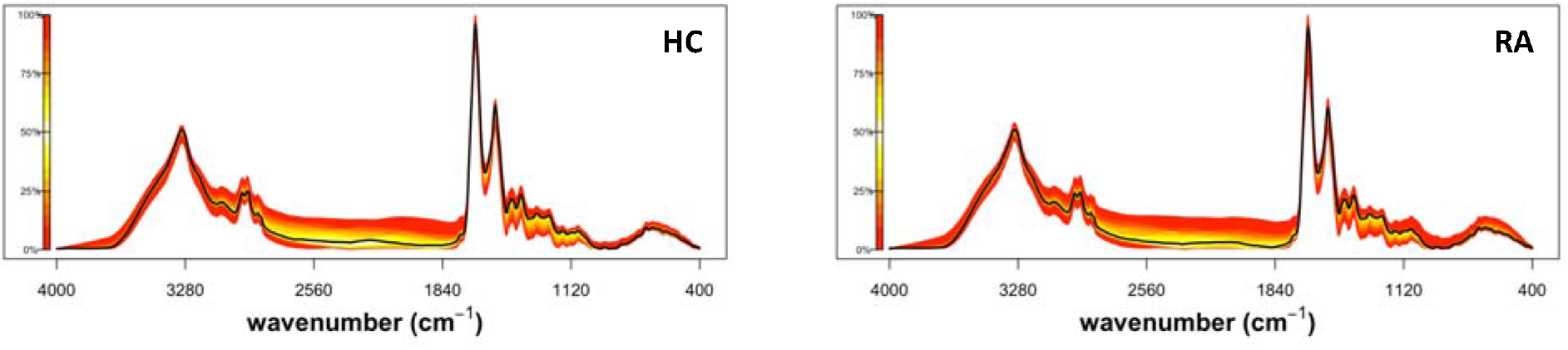
Quantile plot of serum FTIR spectra. Plots depict the median spectral plot (black line) and variation from the median within each cohort, HC (*n*=10) and RA (*n*=20) (yellow to red scale)

### 3.3 Prediction of response to TNF inhibitors using serum metabolomics

We next investigated the potetial of serum metabolomics to predict response to TNF-i therapy in RA. First using ^1^H NMR metabolomics, we saw no separation by PCA (Figure 3A) and univariate analysis found no significant difference between any metabolite bins (data not shown). Similarly, PLS-DA did not show any separation between the RA responders *vs*. non-responders to TNF-i (data not shown) with poor balanced accuracies (55.7%, *p* > 0.1) which were not significantly better than those of NULL modes.

**Figure 3.**
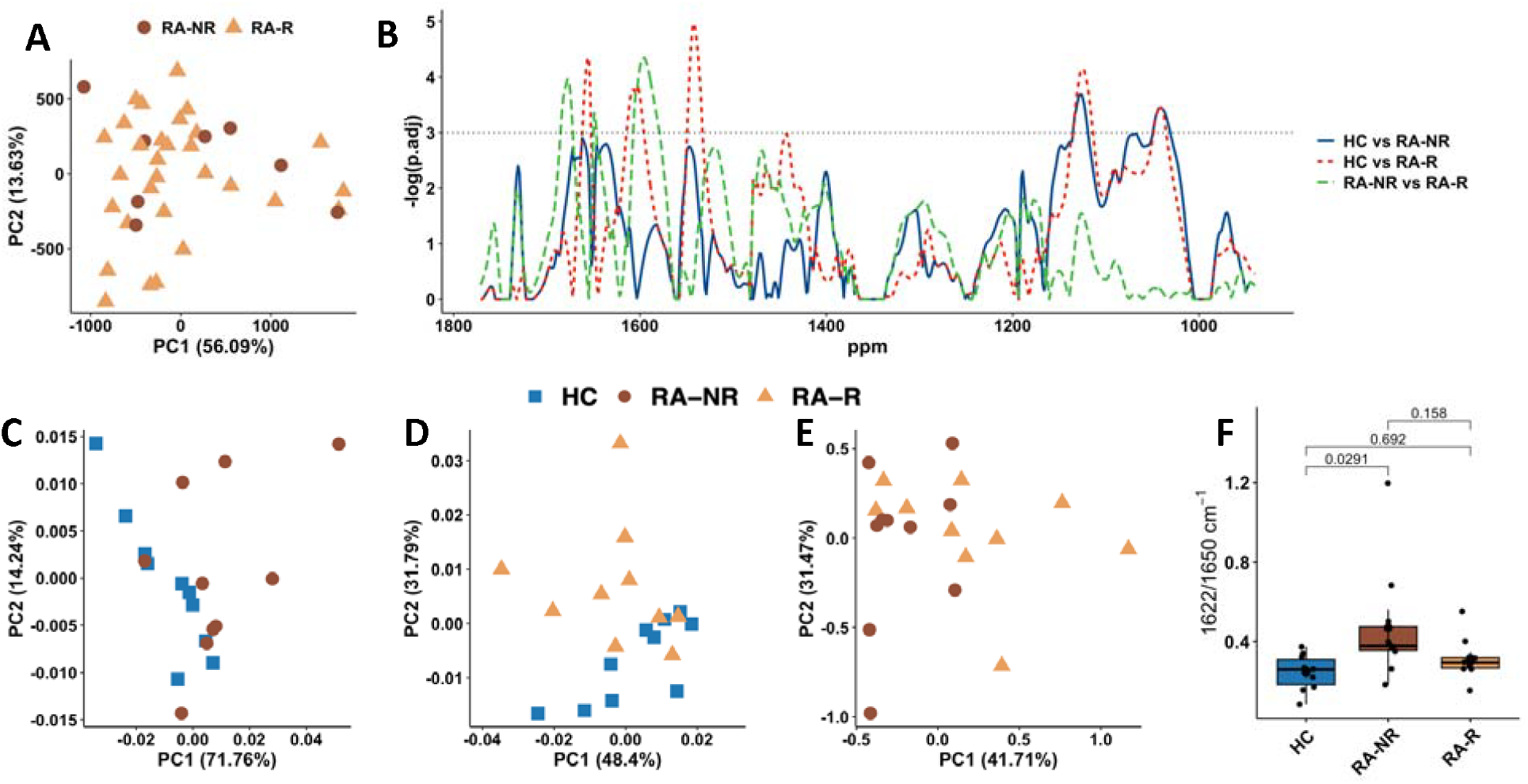
Predicting response to TNFi using NMR and FTIR serum metabolomics. (A) PCA scores plot of 1H NMR analysis of sera from RA TNF-i responders (RA-R, *n=*30) and non-responders (RA-NR, *n=*8). Brackets report the variance explained by the PC. (B) FTIR p-value calculation of HC vs RA TNF-i responders (red), HC vs RA TNF-i non-responders (blue) and RA TNF-i responders vs RA non-responders (green) (*n*=10 each group). The region above by the horizontal black line represents adj.pvalue<0.05. (C) FTIR PCA score plots for HC vs RA TNF-i non-responders, (D) HC vs RA TNF-I responders (E) and RA TNF-i responders vs non-responders. (F) Boxplot showing the ratio of the second derivative of the area integrals of α-helix/β-sheet (1622/1650 cm^-1^) and Tukey post-hoc p-values.

Next we performed a univariate t-test on the FTIR spectra to determine the difference between the RA TNF-i responders and non-responders groups, and against the HC group. When compared against the HC group, both RA groups achieved a significant difference in the 1000-1200 cm^-1^ spectral area, the mixed region of carbohydrates, and nucleic acids (Figure 3B, FDR<0.05). TNF-i responders were significantly different from both HC (FDR<0.05) and TNF-i non-responders (FDR<0.05) in the 1600-1700 cm^-1^ and close to significance in 1450-1500 cm^-1^ spectral regions. PCA of HC and RA TNF-i non-responders serum showed an overlap with clear variance within groups (Figure 3C). HC and RA TNF-i responders showed a partial separation on PC2 (Figure 3D) whereas RA TNF-i responders vs RA TNF-i non-responders were completely overlapping (Figure 3E). We used the second derivative of absorbance intensities ratio at 1622/1651 cm^−1^ to estimate the deformation-induced changes in FTIR spectra and also as a semi quantitative determination of the β-sheet/α-helix content ratio. A significant difference between HC and RA TNF-i non-responders was observed by ANOVA with Tukey’s post-hoc test (Figure 3F, *p*-value=0.029). However no significant difference was observed when comparing RA responders vs HC or RA TNF-i responders vs RA TNF-i non-responders.

PLS-DA models were employed on the fingerprint region of the spectra (wavenumber 940 cm^-1^ to 1771 cm^-1^) to build predictive models for each type of classification (HC *vs*. RA-R, HC *vs*. RA-NR and RA-R *vs*. RA-NR). The results showed that high averaged balanced accuracy achieved by models discriminating HC and RA-R (92.3%,Figure 4A, *p* < 0.001); by contrast, the averaged balanced accuracy was much worse in models discriminating HC and RA-NR (73.1%, Figure 4B *p* < 0.05). For models discriminating RA-R and RA-NR, the averaged balanced accuracy was only 59.7% which is not statistically significant with *p* > 0.1 (data not shown). It is interesting to see that although the results of discriminant analysis of RA-R *vs*. RA-NR was not statistically significant, RA-R and RA-NR have different extent of overlap when compared to the HC group (Figure 4C,D). It is likely that these two group also have subtle differences in their FTIR spectra but the current study does not have sufficient statistical power to detect this, mainly due to very limited number of samples. The distributions of the VIP scores showed that the most important area for classification in both RA groups versus HC was the 1000-1200 cm^-1^ spectral area, the mixed region of carbohydrates, and nucleic acids. The α-helix and β-sheet area for proteins at 1600 to 1700 cm^-1^ was also important in all the models created (Figure 4C,D).

**Figure 4:**
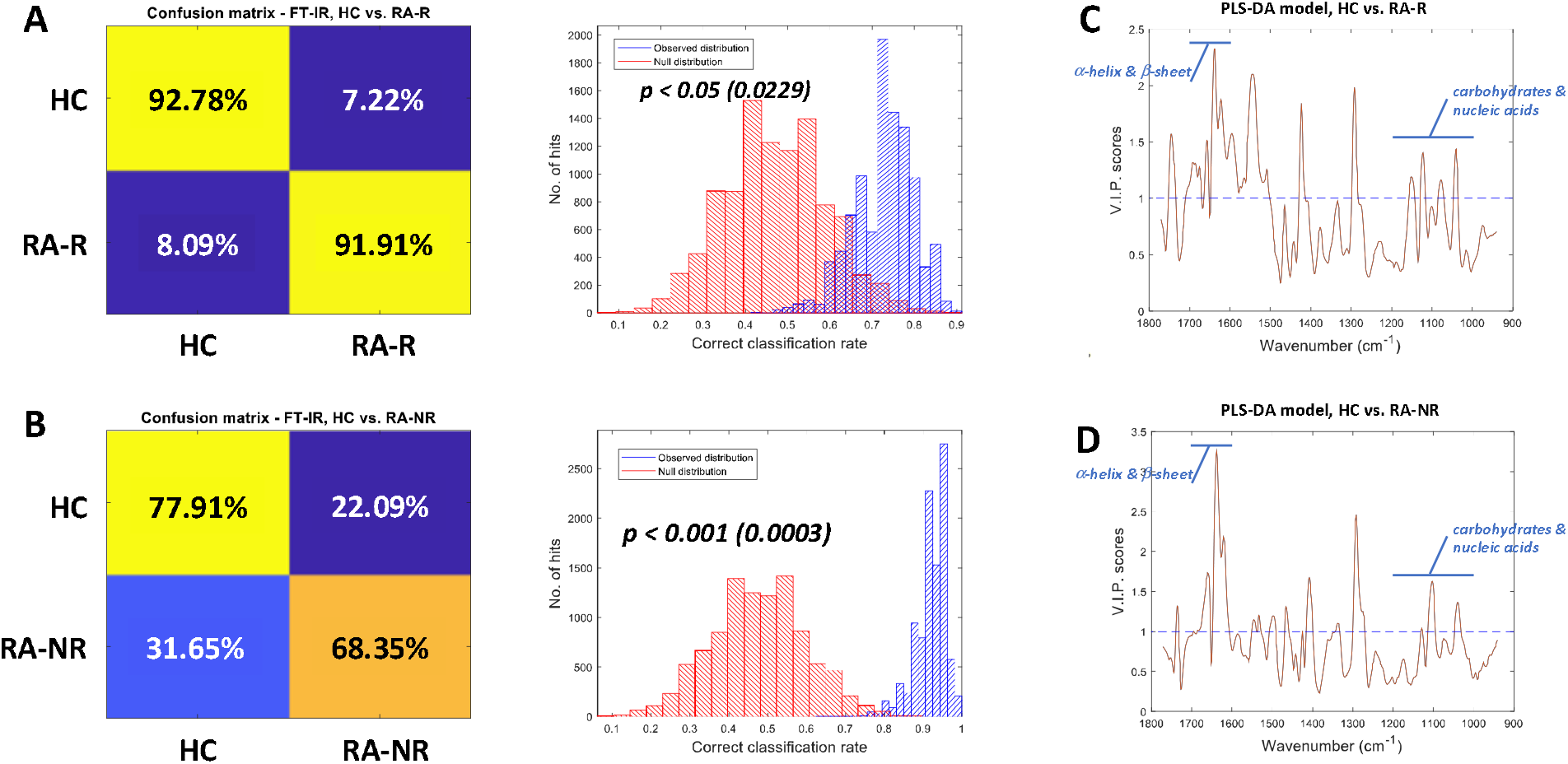
Multivariate analysis of FTIR spectra. (A) Averaged PLS-DA confusion matrix for HC vs RA-R classification and the distributions of performance of observed models (blue) and NULL models(red). (B) Averaged PLS-DA confusion matrix for for HC vs RA-NR classification and distributions of observed models (blue) and NULL models (red). VIP scores plots of PLS-DA model for (C) HC vs RA-R and (D) HC vs RA-NR. Areas of interest relating to carbohydrates and nucleic acids (1000-1200cm^-1^) and a-helix and b-sheet (1600-1700cm^-1^) are indicated in blue.

## 4. Discussion

In this study we have analysed the serum of people with RA and healthy controls to investigate the metabolic composition of this biofluid using two spectroscopy techniques, FTIR and ^1^H-NMR. Previous studies reported significant discriminant properties for comparing HC and RA sera using ^1^H-NMR with valine, isoleucine, lactate, alanine, creatinine and histidine as the main discriminating factors (Dubey et al. 2019; Zabek et al. 2016). Our NMR metabolomics analysis of serum identified acetic acid levels as significantly lower in RA compared to HC, and multivariate modelling also identified 2-hydroxyisovaleric acid, acetoacetic acid, mobile lipids and the amino acids alanine and leucine being the most influential metabolites in PLS-DA models between HC and RA which reached an averaged 83.1% balanced accuracy on blind test sets which was statistically significant compared to the NULL distributions (*p* < 0.05).

Our analysis provides important insight into the metabolic changes associated with inflammation in RA. The most significant difference between RA and HC sera was in the levels of acetic acid, which were lower in RA. Acetic acid is a short chain fatty acid (SCFA) and is produced by the fermentation of dietary fibers by gut microbiota. It is one of several SCFAs that have been implicated in the regulation of immune responses including regulating immune cell activation and differentiation as well as the balance between pro-inflammatory and anti-inflammatory cytokine production (Lin et al. 2023). Lower levels of SCFAs have been observed in the feces of people with RA and in animal models of RA. In antigen-induced arthritis (AIA) mice, supplementation with SCFA decreased arthritis severity and supported regulatory B cell differentiation via the aryl-hydrocarbon receptor (AhR) (Rosser et al. 2020). SCFA can also regulate the differentiation and function of T cells, and promote the differentiation of naïve T cells into regulatory T cells (Tregs), which help in maintaining immune tolerance (Lin et al. 2023). 2-hydroxyisovaleric acid is a breakdown metabolite of branched chain amino acids (BCAA) including leucine, and changes in the levels of these metabolites could reflect increased protein turnover and muscle wasting associated with chronic inflammation (Imai et al. 2024). Increased dietary consumption of BCAA has been associated with development of RA, but not severity of disease (Soleimani Damaneh et al. 2024). Lipid metabolism is also known to be significantly altered in RA (Robinson et al. 2022). People with RA have an increased risk of severe outcomes from cardiovascular disease, despite having low levels of LDL cholesterol and high levels of HDL cholesterol, often described as the RA “lipid paradox” (Choy et al. 2014). DMARD therapies like methotrexate can increase levels of LDL and HDL cholesterol as well as triglyceride levels (Navarro-Millan et al. 2013), and it is worth noting that all RA patients in our study were taking DMARD therapy.

Previous research highlighted the possibility of metabolomics to predict the response to TNF-i therapy in people with RA using urine (Kapoor et al. 2013). Our untargeted analysis was not able to reproduce the same predictive values using serum, reaching a balanced accuracy of 55.7% which was not significantly better than the NULL distribution (*p* > 0.1). Our analysis methods differed in the pre-processing step which could affect the overall results. We annotated the metabolites following MSI guidelines (Sumner et al. 2007) before binning and then carried out univariate and multivariate analysis, whereas the previous study segmented the spectra into increments of 0.005 ppm and annotated the metabolites after the analysis (Kapoor et al. 2013). Another factor may be that the number of biological replicate samples in our study was very limited which results in low statistical power, possibly leading to a false negative result. We additionally analysed the RA sera using FTIR, an affordable, sensitive, specific and user-friendly diagnostic tool to determine if this technology can discriminate people with RA from healthy individuals. An abundance of research shows that IR can characterise and quantify functional groups from the resulting spectra and generates a biochemical fingerprint of the sample (Balan et al. 2019; Huber et al. 2021; Shaw et al. 1999). The particular IR fingerprinting has been thoroughly investigated in the diagnostics discipline and many have shown a promising statistics for application in a clinical environment (Sala et al. 2020) with many proof of concept studies highlighting the ability of FTIR to differentiate between HC and various diseases such as breast, ovarian, bladder, oesophageal and brain cancers, as well as non-malignant diseases such as Alzheimer’s Disease (Backhaus et al. 2010; Butler et al. 2019; Cameron et al. 2019; Gajjar et al. 2013; Hands et al. 2016; Maitra et al. 2019; Ollesch et al. 2014; Paraskevaidi et al. 2017). All these studies have observed these differences in the serum and plasma. These biofluids are present and in high amounts in current biobanks due to the fact that the blood collection is relatively non-invasive and common tests are employed using plasma or serum. The ability to diagnose disease rapidly with high sensitivity and specificity would improve the quality of life and prognosis for patients, in particular those with RA who could receive the most appropriate course of treatment at the developing stages of the disease.

Caveats for analysis of biobanked materials are the variance in sample collection with specific protocols required for serum and plasma collection for both IR and NMR. There are a range of blood collection tubes with a variety of coatings available. These have been analysed by ^1^H-NMR and have shown to have an intrinsic variability between them, including in tubes from the same manufacturer but from different batches (Phelan et al. 2016). For the collection of serum there is no available research on the changes caused by differing vacutainer brands and types. Previous studies have attempted to analyse serum using IR, the first of which analysed 384 serum samples, producing 84% in sensitivity and 88% in specificity in prediction of RA disease to HC (Staib et al. 2001). More recently new studies used ATR-FTIR to compare RA and HC and found significant differences in the serum fingerprint region (Durlik-Popinska et al. 2021; Lechowicz et al. 2016).

In our study we observed that sera from both RA responders and non-responders to TNF-i treatment were different from HC. The 1000-1200 cm^-1^ region was particularly interesting as it was both significantly different in both RA groups compared to HC and is the area with the greatest importance in the PLS-DA projection in the same comparisons. This area predominantly arises due to vibrations from nucleic acids and carbohydrates. Particularly, the appearance of new bands in 1175–1140 cm^−1^ region is believed to be the result of glycosidic linkage formation in polysaccharide, made up of glucose, mannose and fructose (Hong et al. 2021). The overall increase in this region in people with RA could be explained by nutrient factors and their influence on the systemic inflammation. A lower velocity of carbohydrate absorption, presents a negative relationship with CRP, interleukin-6 (IL-6) and TNF-α which are all mediators of inflammation (Ma et al. 2013). Nucleic acids detected as cell free (cf)DNA have also been identified as an important biomarker in autoimmune rheumatic diseases (Duvvuri et al. 2019). In patients with RA, cfDNA levels in peripheral blood and synovial fluid are elevated and have been associated with disease progression (Leon et al. 1977; Leon et al. 1981). Recent studies have demonstrated that NETosis from neutrophils results in a selective extrusion of inflammatory mitochondrial DNA (Hashimoto et al. 2021; Kaplan et al. 2012). The correlation of autoantibodies with the spectral area 1000-1200 cm^-1^ of ATR-FTIR spectra in sera from people with RA (Durlik-Popinska et al. 2021) also suggests how this range is particularly important in the classification of RA serum where autoantibodies are more part of the pathophysiology of the disease.

In FTIR spectra a prediction of α-helix and β-sheet with ∼7% of errors in the prediction is achieved using the absorption at only two wave numbers 1611 and 1652 cm^−1^ (Barth 2007). These peaks have been assigned as vibrational modes of α-helixes and β-sheets respectively in our data to determine differences in HC and both RA groups. The area integral ratio between the two structures proves the robustness as a sensitive and selective screening signature for the discrimination of RA non-responders to HC. Comparing responders to non-responders to TNF-i treatment did not show any significant difference in this ratio. Many studies in RA have already demonstrated the effect of protein mutation to the pathogenesis of this inflammatory disease and others such as inflammatory bowel disease (Huber et al. 2020; Koelink et al. 2014; Kurko et al. 2013) and their manifestation may be the primary reason for the RA non-responder induced α-helix to β-sheet integral ratio difference to HC. For the intra group comparison of RA responders and RA non-responders to TNF-i treatment, FTIR highlighted the 1480-1580 cm^-1^ and 1600-1700 cm^-1^ spectral areas as being significantly different. PLS-DA prediction resulted in 92.3% and 73.1% averaged balanced accuracy for RA responders and RA non-responders respectively. The VIP also pointed to the same areas as being the most important for the PLS-DA model. Both areas are associated with protein content in the samples with the 1480-1580 and 1600-1700 cm^-1^ regions representing amide II and amide I respectively and are characteristic for rheumatoid factor auto-antibodies (Olsztynska-Janus et al. 2012).

A major limitation in this study was our inability to analyse the same clinical samples by both methods due to the limited amount of material available and samples being exhausted by multiple experiments and technical replicates. Furthermore, for a more robust statistical and multivariate analysis more samples would be needed to increase the reliability and statistical power of the NMR and FTIR findings. Finally the donors were not required to fast before the blood collection and samples were obtained at different times in the day which would affect the NMR small metabolite abundances and increase variances between patients in the same group. Nevertheless, we believe this pilot study has shown the value in analysing human serum by NMR and FTIR and has tentatively shown that it is possible to discriminate sera from people with RA from HC that would benefit from further investigation in a larger and more well-defined clinical cohort.

In conclusion, this work has demonstrated the clinical usability of FTIR in the diagnosis of RA. This technique offers a simple, cost-effective solution for routine screening using only serum which is readily available and can be ontained in the clinic and can be used alongside routine blood tests. The alteration of unique spectral markers in the serum may be an important diagnostic tool for the future early confident detection of RA in combination with other existing criteria such as X-ray images highlighting erosions, auto-antibody titres and non-specific markers of inflammation (ESR and CRP). Furthermore it could increase the ability to stratify RA patients as responders to or non-responders to treatments which would be an advantageous asset in the treatment process of RA.

## Data Availability

All data produced in the present study are available upon reasonable request to the authors.

## 6 Acknowledgements

We would like to thank clinical and laboratory staff within the rheumatology department at the local hospital Trusts for help collecting serum samples.

## 9. Author Contributions

Conceptualisation HLW, MMP, RG; Methodology MMP, RG, YX, CL, MFA; Formal analysis MFA, YX, SF, HLW; Investigation MFA, CL; Visualisation MFA, YX, MMP, HLW; Data curation, MMP, CL, RG; Writing the manuscript MFA, YX, SF, HLW, MP; Revising the manuscript CL, RG. All authors have read and agreed the final version of the manuscript.

## 10. Author Contributions

All data produced in the present study are available upon reasonable request to the authors.

---

**Supplementary Table 1:** Discriminatory Infrared spectral bands, with biomolecular assignments and their bond vibrations, adapted from (Ghimire et al. 2020; Ramalingam et al. 2014). vas = asymmetric stretching vibration, vs = symmetric stretching vibration, v = v stretching vibration

| Band (cm<sup>-1</sup>) | Assignment and vibrations |
| --- | --- |
| 900-1158 | Carbohydrates (Glucose, Mannose, Fructose) and nucleic acids (Deoxyribose/Ribose DNA, RNA): C-O, C-C stretch, C-H bends, Endocyclic C-O-C vibration and, vs(PO<sup>2-</sup>) |
| 1208-1244 | Amide III, vas(PO<sup>2-</sup>) |
| 1317-1382 | Collagen: CH<sub>2</sub> wagging, the vibration of α, and β anamor |
| 1420-1430 | Polysaccharides, vs (COO-), (CH<sub>2</sub>) |
| 1480-1580 | Amide II of proteins: (α-helical, β-pleated sheet, unordered conformation structures), δ(N-H), v(C-N) |
| 1600-1700 | Amide I of proteins: (α-helical, β-pleated sheet, β-turns, random coils, and sidechain, β (anti-□+turn) structures), v(C=O), v(C-N), CNN |
| 1720-1750 | Lipids C=O stretching |

